# A Case for Estradiol: Younger Brains in Women with Earlier Menarche and Later Menopause

**DOI:** 10.1101/2024.04.20.24306043

**Authors:** Eileen Luders, Inger Sundström Poromaa, Claudia Barth, Christian Gaser

## Abstract

The transition to menopause is marked by a gradual decrease of estradiol. At the same time, the risk of dementia increases around menopause and it stands to reason that estradiol (or the lack thereof) plays a significant role for the development of dementia and other age-related neuropathologies. Here we investigated if there is a link between brain aging and estradiol-associated events, such as menarche and menopause. For this purpose, we applied a well-validated machine learning approach in a sample of 1,006 postmenopausal women who were scanned twice approximately two years apart. We observed less brain aging in women with an earlier menarche, a later menopause, and a longer reproductive span (i.e., the time interval between menarche and menopause). These effects were evident both cross-sectionally and longitudinally, which supports the notion that estradiol might contribute to brain preservation. However, more research is required as effects were small and no direct measures of estradiol were obtained in the current study.

## Introduction

Estradiol is the most potent and prevalent form of estrogen during the reproductive life of a woman^1^. Generally speaking, estradiol levels start increasing just before the first menstrual period (menarche) and then plateau on a high level until they start decreasing during perimenopause. After the final menstrual period (i.e., menopause) estradiol levels decrease further and eventually reach plateauing low levels during postmenopause^2^. The risk for dementia in women is known to increase around menopause^3-6^ and thus it stands to reason that estradiol plays a significant role for the development of dementia and other age-related neuropathologies. However, scientific studies focused on specific phases (e.g., menarche, pregnancy, menopause) or interventions (oral contraceptives, estrogen modulation therapy, estrogen replacement therapy) paint a rather complex picture, with no consistent evidence that more (estradiol) is always beneficial^4,7-22^. For example, with particular respect to menarche and menopause, both early onset and late onset have been shown to increase the risk for dementia or to be positively associated with brain aging and cognitive functioning^4,7-14,18-20^.

To further advance this field of research, the current study set out to determine if there is a link between a woman’s estimated brain age (a biological marker of brain health^23^) and the reproductive span (i.e., the interval between menarche and menopause when estradiol levels are high). If a lack of estradiol is among the driving factors for diminished brain health later in life, brain age and reproductive span should be inversely related (negative correlation). To be able to relate our findings to others in the literature^7-9^ and to provide a frame of reference for future studies, we additionally investigated if there is a significant link between estimated brain age and the age at menarche as well as the age at menopause. Assuming a neuroprotective effect of estradiol, we expected that a lower brain age would be linked to an earlier menarche (positive correlation) and to a later menopause (negative correlation). Importantly, our study comprises both cross-sectional and longitudinal components, with follow-up data acquired approximately two years after the initial brain scan. This constitutes a critical extension to existing studies from the UK biobank with a related focus as those studies are solely cross-sectional in nature^8,9,11,17,18,19^.

Our study was conducted in a sample of 1,006 postmenopausal women whose brain ages were estimated using structural brain images and a well-validated high-dimensional pattern recognition approach, as detailed elsewhere ^24,25^. Briefly, the difference between the estimated brain age and the chronological age yields a so-called brain age gap estimate (BrainAGE) in years. The BrainAGE index is negative if a brain is estimated younger than its chronological age; it is positive if a brain is estimated older than its chronological age. For example, a 50-year-old woman with a BrainAGE index of -3 years shows the aging pattern of a 47-year-old. The BrainAGE algorithm has been shown to be robust and reliable across datasets, age-ranges, and scanner types^24,26^; it has been successfully applied in a wide range of studies^24,25,27-29^ including those capturing hormonal changes in women^30,31^. Moreover, the BrainAGE index has been demonstrated to work as a predictor of dementia as well as age-related cognitive decline^27,32^.

## Results

### Main Analysis

As shown in **Figure 1** (left), our cross-sectional analyses revealed a significant negative association between BrainAGE and the reproductive span. In other words, brains of women with longer reproductive spans were estimated younger than brains of women with shorter reproductive spans. As also shown in **Figure 1** (right), there was a significant positive association between BrainAGE and age at menarche (i.e., the earlier the menarche, the younger the brain) and a significant negative association between BrainAGE and age at menopause (i.e., the later the menopause, the younger the brain). Statistics are provided in **Table 1**, including the slopes of the regression which indicate different rates of change for menarche and menopause (0.32 and -0.10, respectively): More specifically, for each year younger at menarche, brains are estimated 0.32 years younger (which corresponds to 3.2 years younger for each 10 years). In contrast, for each year older at menopause, brains are estimated 0.1 year younger (which corresponds to 1 year younger for each 10 years).

**Table 1.** Associations with BrainAGE at the initial brain scan.

| | $R^2$ | Correlation (r) | Significance (p) | Slope |
| --- | --- | --- | --- | --- |
| Reproductive Span | 0.01 | -0.11 | <0.001 | -0.11 |
| Age at Menarche | 0.02 | 0.14 | <0.001 | 0.32 |
| Age at Menopause | 0.01 | -0.09 | <0.005 | -0.10 |

**Figure 1:**
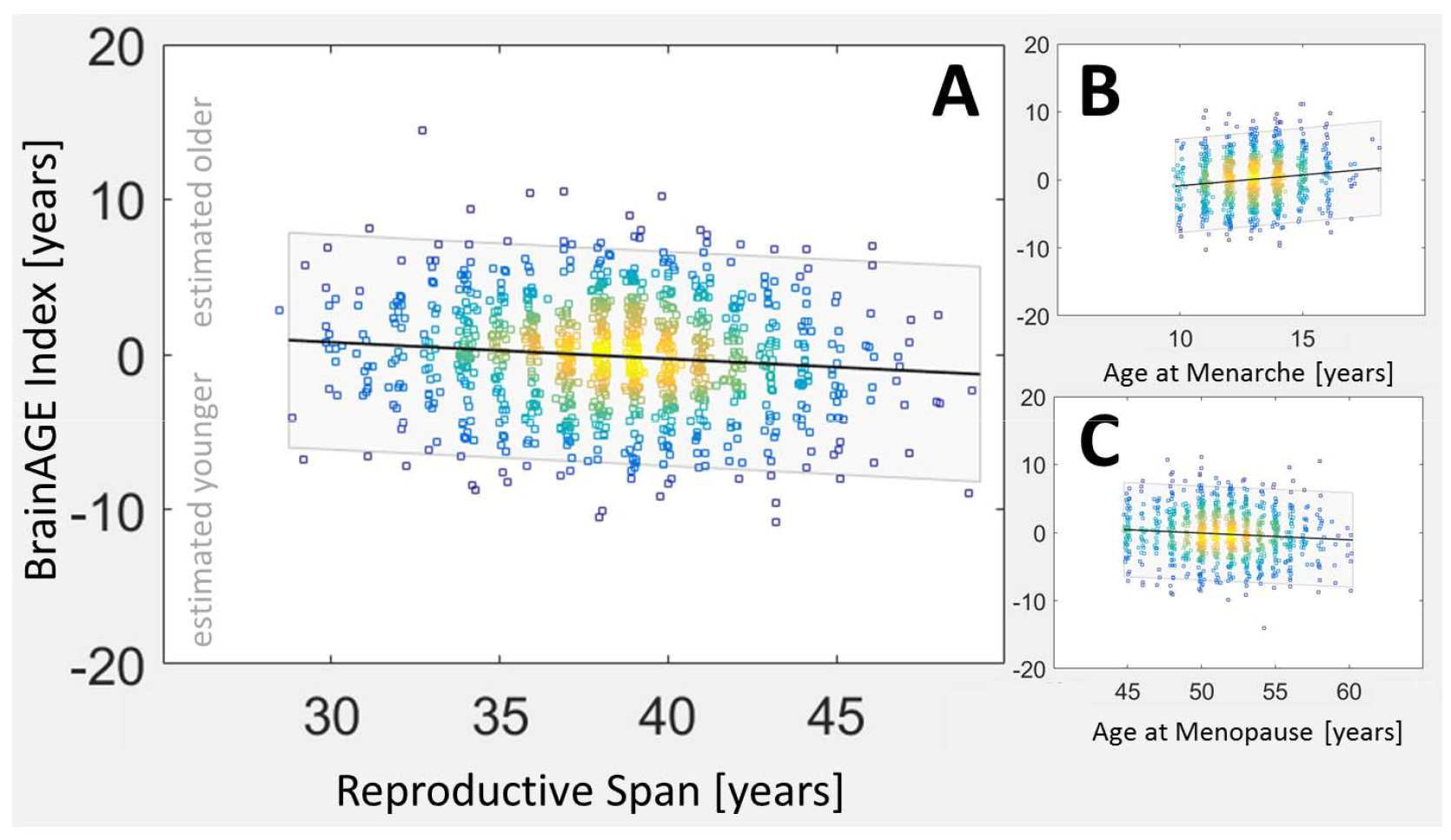
Correlations with BrainAGE at the initial brain scan. The x-axes show the reproductive span (age, respectively) in years. Of note, age in the UK Biobank has been rounded to the year, so we added a small random jitter to the x-axes to give a better overview about the age distribution. The y-axes show the BrainAGE index in years, with negative values indicating that brains are estimated younger than their chronological age and positive values indicating that brains are estimated older than their chronological age. Panel A displays a negative link between the BrainAGE index and the reproductive span (the longer the reproductive span, the younger the estimated brain age). Panel B displays a positive link between the BrainAGE index and the age at menarche (the earlier the onset of menarche, the younger the estimated brain age). Panel C displays a negative link between the BrainAGE index and the age at menopause (the later the onset of menopause, the younger the estimated brain age). Hot colors in the density plot indicate a larger overlay of measures; cool colors indicate a smaller overlay. The shaded band is the 95% confidence interval.

As shown in **Figure 2** and **Table 2**, our longitudinal findings confirm the observed cross-sectional relationships. More specifically, Δ BrainAGE was negatively linked to reproductive span and menopause, and positively linked to age at menarche. All associations were significant. Moreover, the slopes of the regression are still somewhat different for menarche and menopause (0.08 and -0.06, respectively), albeit more similar than in the cross-sectional analysis: For each year younger at menarche, brains are estimated 0.08 years younger (0.8 years over 10 years), whereas for each year older at menopause, brains are estimated 0.06 years younger (0.6 years over 10 years).

**Table 2.**
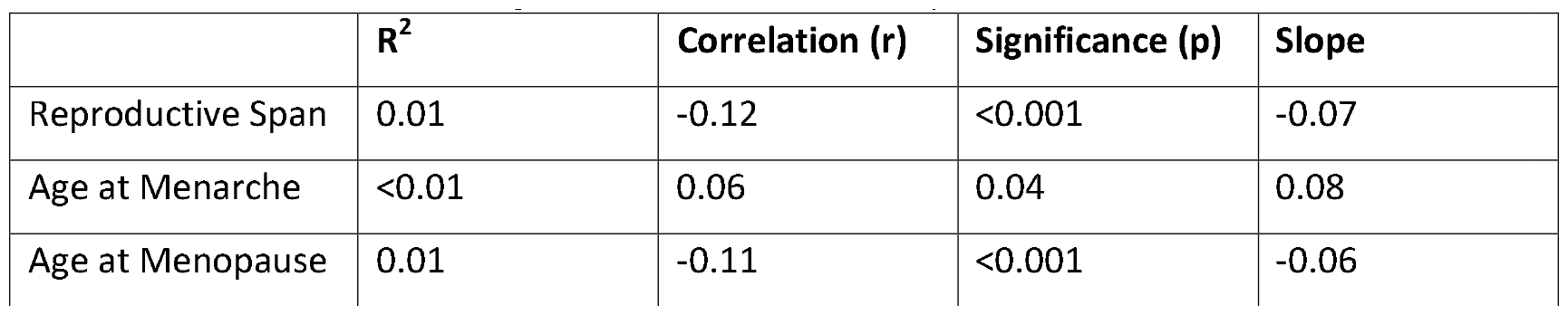
Associations with changes in BrainAGE over 2.35 years.

| | $R^2$ | Correlation (r) | Significance (p) | Slope |
| --- | --- | --- | --- | --- |
| Reproductive Span | 0.01 | -0.12 | <0.001 | -0.07 |
| Age at Menarche | <0.01 | 0.06 | 0.04 | 0.08 |
| Age at Menopause | 0.01 | -0.11 | <0.001 | -0.06 |

**Figure 2:**
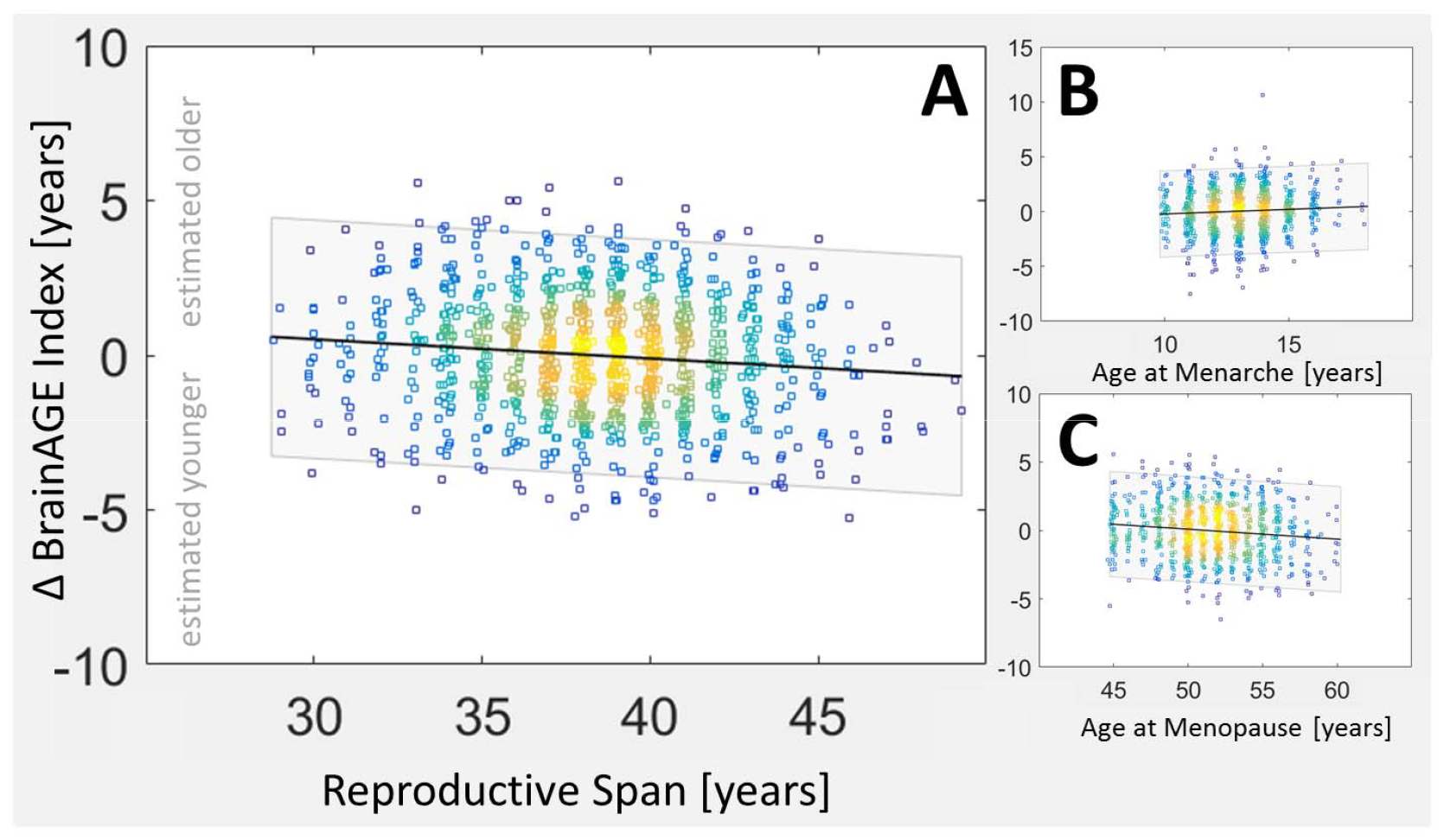
Correlations with BrainAGE over 2.35 years (Δ BrainAGE). Panel A displays a negative link between the BrainAGE index and reproductive span (the longer the reproductive span, the smaller the estimated brain age). Panel B displays a positive link between the BrainAGE index and the age at menarche (the earlier the onset of menarche, the younger the estimated brain age). Panel C displays a negative link between the BrainAGE index and the age at menopause (the later the onset of menopause, the younger the estimated brain age). Hot colors in the density plot indicate a larger overlay of measures; cool colors indicate a smaller overlay. The shaded band is the 95% confidence interval.

### Sensitivity Analysis

The results described above remained stable when removing the variance associated with the number of live births, hormone replacement therapy, hysterectomy, bilateral oophorectomy, body mass index, diastolic and systolic blood pressure, diabetes, education, income, and a composite lifestyle factor. In other words, when examining the association between BrainAGE and reproductive span, we observed a negative association. Likewise, there was a positive association between BrainAGE and age at menarche and a negative association between BrainAGE and age at menopause. The effects were significant both for the cross-sectional analyses (see **Supplemental Table 1**) and the longitudinal analyses (see **Supplemental Table 2**).

## Discussion

Here we assessed links between estimated brain age and milestones in a woman’s reproductive life in a well-powered sample of more than a thousand postmenopausal women. We detected less brain aging in women with longer reproductive spans, earlier menarche, and later menopause.

### Correspondence with Previous Findings

Our findings are in line with the outcomes of other studies suggesting a longer reproductive span ^8,10,13^, an earlier menarche^13,14^, as well as a later menopause ^4,8,10-12^ to be associated with a lower risk of developing dementia or better retained cognitive function. Furthermore, given that the BrainAGE index is based on the weighted distribution of gray and whiter matter tissue in the brain, our findings are also in agreement with reports of lower brain volumes as well as higher rates of brain tissue loss during menopause compared to premenopause or in postmenopausal women compared to premenopausal women^33-35^. In addition, our findings agree with observed effects across the menstrual cycle linking high estradiol levels at ovulation to lower BrainAGE estimates^30^. Altogether, the outcomes of our study seem to suggest that estradiol contributes to brain health, which is in agreement with other studies reporting positive effects of estradiol on brain health and cognition within the framework of aging and/or menopausal hormone therapy^36-40^.

### Menarche versus Menopause

In the present study, both an earlier menarche and a later menopause were significantly associated with less brain aging. However, aside from the mere direction of the relationship, menarche and menopause also differ with respect to the strength of their relationship with age (which is reflected in the correlation coefficient) and their rate of change with age (which is reflected in the slope of the regression line). This might indicate somewhat different underlying biological mechanisms and/or confounds. For example, during menopause, in addition to decreasing levels of estradiol, increasing levels of follicle-stimulating hormones may cause an accelerated deposition of amyloid-β and Tau^41^, which enhances brain atrophy. Moreover, menopause is marked by disadvantageous alterations in cytokine and T cell profiles^42^, which are linked to an enhanced inflammation.

### Possible Implications

Given that estradiol levels start decreasing during perimenopause and further decrease after menopause, our findings may explain why the risk for dementia in women is known to increase around menopause^3-6^ and why there is an increased age-independent prevalence of Alzheimer’s disease in women compared to men^40^. Moreover, our findings seem to support to the concept of the “window of opportunity”, spanning the years leading up to menopause to the years immediately after menopause, where health interventions (e.g., menopausal hormone treatment) may combat the increased risk for Alzheimer’s disease in some women^5,43-45^. However, at this point, all of this is conjecture. In fact, several large-scale projects have investigated the effects of menopausal hormone treatment on cognitive function and Alzheimer’s risk, but results are inconclusive (potentially relevant modulators of treatment outcomes are discussed here^36,46-51^).

### A Word of Caution

Our findings seem promising in the framework of prevention and intervention. However, the effect sizes for the observed associations between estimated brain age and reproductive span, age at menarche, and age at menopause were small. This raises the question of whether the apparent impact of estradiol is clinically meaningful. Perhaps, equally relevant, we wish to emphasize that the study did not measure estradiol directly. Therefore, further research is required, the more so as links between estradiol and brain aging are rather complex as indicated by the outcomes of other studies. For example, it was reported that, compared to no exposure or no dose, exposure to low concentrations of estradiol or low doses of estrogen enhanced neuronal survival and increased anti-inflammatory markers (i.e., positive links), while exposure to high concentrations of estradiol as well as high doses of estrogen had the opposite effect (i.e., negative links)^21,22^. Another study reported U-shaped curves suggesting that both early and late menarche are associated with an increased risk for dementia (i.e., positive and negative links)^8^. And yet another study reported either negative links or missing links between age at menarche and brain aging depending on the potential confounds accounted for^7^. Interestingly, this latter study also reported that, in carriers of the apolipoprotein E type 4 allele (APOE e4), higher levels of estradiol at menopause were associated with increased brain aging (positive link). In contrast, in non-carriers, higher levels of estradiol at menopause were associated with decreased brain aging (negative link)^7^.

## Conclusion

Our study revealed less brain aging in women with a larger reproductive span, earlier menarche, and later menopause. Thus, sex hormones – potentially estradiol – may contribute to brain health. However, follow-up research is required because effects in the current study were small, estradiol was not directly examined, and female brain health may be modulated by other factors than estradiol^2,52,53^. Moreover, to paint a more complete picture and expand an understudied field research, future research focussing on specific time frames surrounding menopause, such as perimenopause (i.e., the time preceding the final menstrual period) or early postmenopause (e.g., the initial year after menopause) versus late menopause (e.g., ten years after menopause) would be desirable. Last but not least, the UK Biobank is biased towards healthy^54^ and more socioeconomically privileged individuals^54^ with a predominant white ethnic background^54^, which affects the generalizability of the findings.

## Methods

## Sample

The study is based on a carefully selected sample of postmenopausal women from the UK Biobank (https://www.ukbiobank.ac.uk/) which was accessed under application number #41655. The UK Biobank is a biomedical database and research resource that contains genetic, lifestyle and health information from half a million people. In the UK Biobank cohort, 94.6% of participants are of white ethnicity^54^. For general ethnic information, see https://biobank.ctsu.ox.ac.uk/crystal/field.cgi?id=21000; for ethnic information on all women with available longitudinal data, see **Supplemental Table 3**. The UK Biobank holds the ethical approval from the North West Multi-Centre Research Ethics Committee (MREC) and is in possession of the informed consents. Written informed consent was obtained from all participants.

**Table 3.** Sample characteristics.

| Variable | Descriptive Statistics |
| --- | --- |
| age at the initial brain scan* | 63.20 ± 6.42 years |
| age at the follow-up brain scan* | 65.54 ± 6.37 years |
| age at menarche* | 13.02 ± 1.53 years |
| age at menopause* | 51.41 ± 3.23 years |
| reproductive span* | 38.39 ± 3.55 years |
| number of live births* | 1.75 ± 1.16 |
| number of women with hormone replacement therapy | yes: 306 \| no: 700 |
| number of women with hysterectomy | yes: 48 \| no: 958 |
| number of women with bilateral oophorectomy | yes: 37 \| no: 969 |

\*statistics provided as mean ± standard deviation

Exclusion criteria for the current study were pre-existing neurological or psychiatric diagnoses as per UK Biobank data fields #41202-0.0 to #41202-0.78. Inclusion criteria for the current study were women with available longitudinal data as well as information on age at menarche and age at menopause. In addition, to further increase the homogeneity of the sample, we excluded women whose age at menarche was younger than 10 or older than 18, or whose age at menopause was younger than 45 or older than 60. This resulted in a final sample size of 1,006 women. **Figure 3** summarizes the steps related to the sample selection; **Table 3** provides information on the final sample.

**Figure 3.**
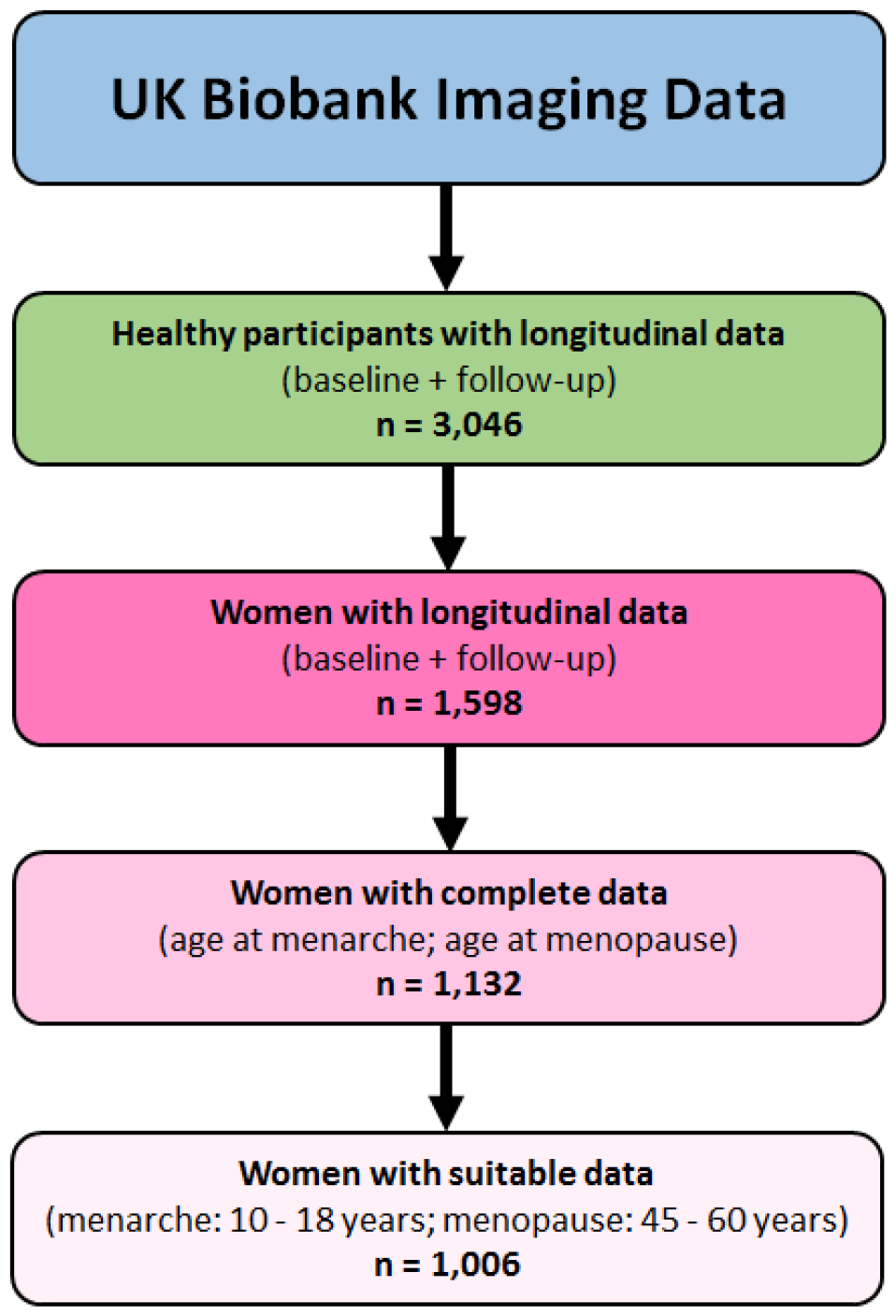
Flowchart of sample selection.

## Image Acquisition and Processing

For each woman, one initial brain scan and one follow-up brain scan – approximately two years apart (mean ± SD: 2.35 ± 6.12 years) – were obtained after menopause. Brain images were acquired on a 3 Tesla Siemens Skyra scanner using a 32-channel head coil, as described elsewhere^55^ (see also: http://biobank.ctsu.ox.ac.uk/crystal/crystal/docs/bmri_V4_23092014.pdf). Using the T1-weighted images, we applied a number of processing routines implemented in the CAT12 toolbox^56^ (version 12.8) that resulted in bias-corrected, spatially normalized, and tissue-classified brain images, as detailed elsewhere^24,31^. The affinely normalized gray and white matter partitions were then smoothed using a 4 and 8 mm full-width-at-half-maximum (FWHM) Gaussian kernel, and image resolution was set to 4 and 8 mm. For further data reduction, we applied a principal component analysis (PCA) using singular value decomposition to all the models using n–1 PCA components (n = minimum of voxel number or sample size). For the estimation of the BrainAGE index, we employed a Gaussian Process Regression (GPR) that uses a linear covariance function, a constant mean function, and a Gaussian likelihood function; hyperparameters were set to 100 for the constant mean function and to -1 for the likelihood function^57^. As training data, we selected 3,046 individuals from the UK Biobank where two time points were available and applied a 10-fold validation approach separately for the initial and follow-up brain scan. To estimate the individual brain ages, eight models based on the aforementioned sets of images (i.e., gray matter/white matter, 4 mm/8 mm Gaussian kernel, and 4 mm/8 mm image resolution) were combined using a general linear model where the weights of the models were derived by maximizing the variance to the parameter of interest (e.g., menopause). The difference between the resulting estimated brain age and the chronological age was then calculated as the BrainAGE index (in years).

## Statistical Analyses

After computing the BrainAGE index for all 1,006 women at initial and follow-up scan, we first removed the linear age trend that is typically seen in BrainAGE estimation. Then, we conducted two analysis streams using linear regressions, one cross-sectional and one longitudinal. For the cross-sectional stream, we tested if there is a significant link between the BrainAGE index at the initial brain scan and the reproductive span. In addition, we tested if there is a significant link between the BrainAGE index at the initial brain scan and the age at menarche as well as the age at menopause. For the longitudinal stream, we first subtracted the BrainAGE index at the initial brain scan from the BrainAGE index at the follow-up brain scan, which resulted in a Δ BrainAGE index for each individual. This method, often referred to as “change score” analysis, produces statistical results that are comparable to those resulting from a repeated-measures ANOVA with two time points. Using the Δ BrainAGE index, we then tested for significant links with the reproductive span, the age at menarche, and the age at menopause. For all analyses, alpha was set at 0.05 (two-tailed).

## Sensitivity Analyses

To determine if our results remain stable and significant when accounting for potential confounds known to affect brain health, we repeated the aforementioned cross-sectional and longitudinal analyses for reproductive span, menarche, and menopause using additional parameters. More specifically, we removed the variance associated with the number of live births^58^ (UK Biobank data field #2734), hormone replacement therapy^7^ (#2814), hysterectomy^59^ (#3591), bilateral oophorectomy^59^ (#2834), body mass index^60^ (#21001), diastolic and systolic blood pressure^61^ (#4079 and #4080), diabetes^62^ (#2443), education^63^ (#6138), income^64^ (#738), and a composite lifestyle factor^65^. The latter was expressed as a general lifestyle score that was calculated based on a number of factors (see Supplemental Table 4), known to increase / decrease the risk of adverse cardiovascular events. Since not all women had information on all potential confounds, we applied an imputation method using the Matlab function ‘fillmissing’. That is, missing entries were replaced with the corresponding values from the nearest neighbor rows, calculated based on the pairwise Euclidean distance between rows. Imputation was applied to up to 295 women, depending on the potential confound.

## Acknowledgments

EL was supported by the Swedish Collegium for Advanced Study (SCAS) and Erling-Persson Family Foundation. The data of the UK Biobank were accessed under application number #41655. The article was composed using the STROBE cohort checklist^66^.

## Data Availability

Publicly available datasets were analyzed in this study and are available here: https://www.ukbiobank.ac.uk

## Code Availability

The code for processing the data is available here: https://github.com/ChristianGaser/cat12

The code for estimating BrainAGE is available here: https://github.com/ChristianGaser/BrainAGE

## Supplemental Material

**Supplemental Table 1.**
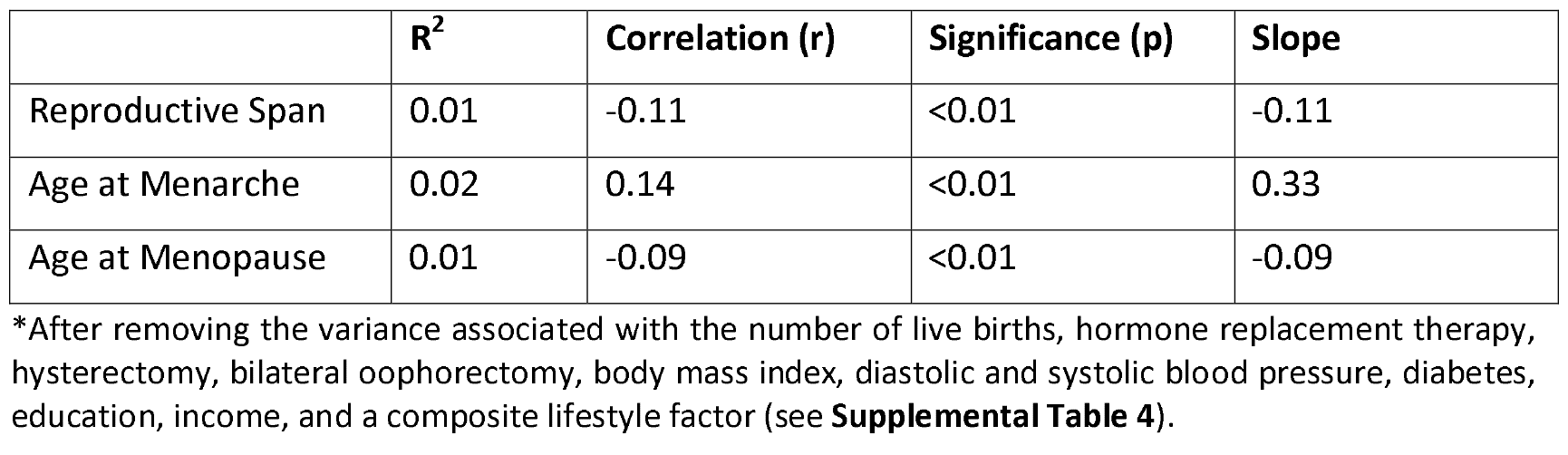
Associations with BrainAGE at the initial brain scan (adjusted model^*^)

|  | R<sup>2</sup> | Correlation (r) | Significance (p) | Slope |
| --- | --- | --- | --- | --- |
| Reproductive Span | 0.01 | -0.11 | <0.01 | -0.11 |
| Age at Menarche | 0.02 | 0.14 | <0.01 | 0.33 |
| Age at Menopause | 0.01 | -0.09 | <0.01 | -0.09 |

\*After removing the variance associated with the number of live births, hormone replacement therapy, hysterectomy, bilateral oophorectomy, body mass index, diastolic and systolic blood pressure, diabetes, education, income, and a composite lifestyle factor (see **Supplemental Table 4**).

**Supplemental Table 2.**
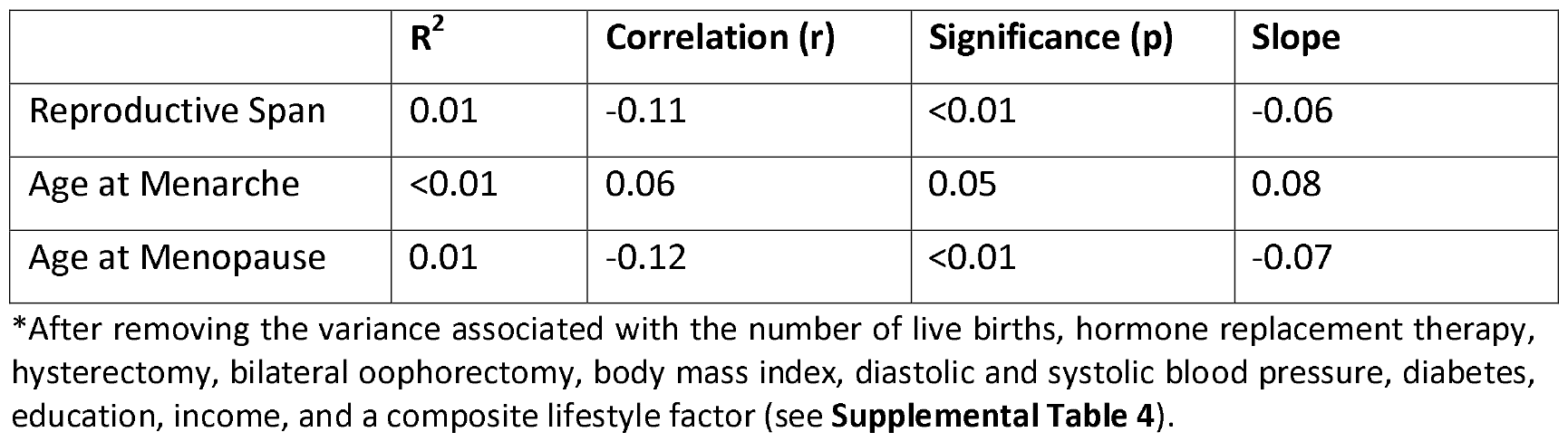
Associations with BrainAGE over 2.35 years (adjusted model^*^)

**Supplemental Table 3.** Ethnic background of women with longitudinal MRI data (n=1,598)

| Ethnicity | Number of Women |
| --- | --- |
| British | 1497 |
| Any other white background | 42 |
| Irish | 25 |
| Chinese | 9 |
| Other ethnic group | 5 |
| Caribbean | 5 |
| Indian | 3 |
| Pakistani | 3 |
| African | 3 |
| Any other mixed background | 3 |
| White and Black Caribbean | 2 |
| Any other Asian background | 1 |
| White and Black African | 0 |

Information on ethnicity was collected based on self-reports using a touchscreen questionnaire at the UK Biobank Assessment Centre.

**Supplemental Table 4.** A general lifestyle score was calculated based on 16 variables.

| **Variables** | **UK Biobank Data Field Number** |
| --- | --- |
| 1. Time spend watching TV | #1070 |
| 2. Sleep duration | #1160 |
| 3. Current tobacco smoking | #1239 |
| 4. Past tobacco smoking | #1249 |
| 5. Cooked vegetable intake | #1289 |
| 6. Salad / raw vegetable intake | #1299 |
| 7. Fresh fruit intake | #1309 |
| 8. Dried fruit intake | #1319 |
| 9. Oily fish intake | #1329 |
| 10. Processed meat intake | #1349 |
| 11. Beef intake | #1369 |
| 12. Lamb mutton intake | #1379 |
| 13. Pork intake | #1389 |
| 14. Alcohol intake frequency | #1558 |
| 15. Moderate activity | #884 |
| 16. Vigorous activity | #904 |

